# Genomic risk scores for juvenile idiopathic arthritis and its subtypes

**DOI:** 10.1101/2020.02.20.20025924

**Authors:** Rodrigo Cánovas, Joanna Cobb, Marta Brozynska, John Bowes, Yun R. Li, Samantha L Smith, Hakon Hakonarson, Wendy Thomson, Justine Ellis, Gad Abraham, Jane Munro, Michael Inouye

## Abstract

**Aims:** Juvenile idiopathic arthritis (JIA) is an autoimmune disease and a common cause of chronic disability in children. Diagnosis of JIA is based purely on clinical symptoms, leading to treatment delays. Despite JIA having substantial heritability, the construction of genomic risk scores (GRSs) to aid or expedite diagnosis has not been assessed. Here, we generate GRSs for JIA and its subtypes and evaluate their performance.

**Methods:** We examined three case/control cohorts (UK, US, and Australia) with genome-wide single nucleotide polymorphism (SNP) genotypes. We trained GRSs for JIA and its subtypes using lasso-penalised linear models in cross-validation on the UK cohort, and externally tested in the Australian and US cohorts.

**Results:** The JIA GRS alone achieved cross-validated AUC=0.670 in the UK cohort and externally validated AUCs of 0.657 and 0.671 in US-based and Australian cohorts, respectively. In logistic regression of case/control status, the corresponding odds ratios per standard deviation (s.d.) of GRS were 1.831 [1.685-1.991] and 2.008 [1.731-2.345], and were unattenuated by adjustment for sex or the top 10 genetic principal components. Extending our analysis to JIA subtypes revealed that enthesitis-related JIA had both the longest time-to-referral and the subtype GRS with the strongest predictive capacity overall across datasets: AUCs 0.80 in UK; 0.83 Australian; 0.69 US-based. The particularly common oligoarthritis JIA subtype also had a subtype GRS outperformed those for JIA overall, with AUCs of 0.71, 0.75 and 0.77, respectively.

**Conclusions:** A genomic risk score for JIA has potential to augment purely clinical JIA diagnosis protocols, prioritising higher-risk individuals for follow-up and treatment. Consistent with JIA heterogeneity, subtype-specific GRSs showed particularly high performance for enthesitis-related and oligoarthritis JIA.

## Introduction

Juvenile idiopathic arthritis (JIA) is an autoimmune disease that comprises all form of arthritis arising before the age of 16 years and persisting for more than 6 weeks^1^. JIA has a significant impact on quality of life, physical function, and future development, and its prevalence is estimated at 0.07–4 per thousand children of European descent^2–4^. The International League of Associations of Rheumatology (ILAR) classification system recognises seven subtypes of JIA based on the number of joints affected, age of onset, and other features^5^. The aetiology of JIA is not well understood and its diagnosis remains purely clinical with no sensitive or specific tests available to assist clinicians in making the diagnosis.

Early diagnosis and treatment of JIA is critical as delays increase the risk of prolonged and more difficult to control diseases and consequently poorer long-term outcomes^6–9^. However, in most cases, general practitioners, emergency medicine physicians, and paediatricians have limited experience in recognising and diagnosing JIA. This might affect time to diagnosis in two ways (**Figure 1**). Firstly, JIA may not be suspected in a symptomatic child, and secondly, children with generalised symptoms may be incorrectly referred to paediatric rheumatologists for management^10–12^, putting undue pressure on clinics and waiting lists. For example, a study in Australia^13^ found that the average time from parent reported symptom onset to diagnosis of JIA by a paediatric rheumatologist was 7 months^14^. Therefore, there is an urgent need for tools that assist clinicians in assessing children with musculoskeletal symptoms who may be JIA cases, and thus reduce complications of the disease and poor long-term health outcomes.

**Figure 1:**
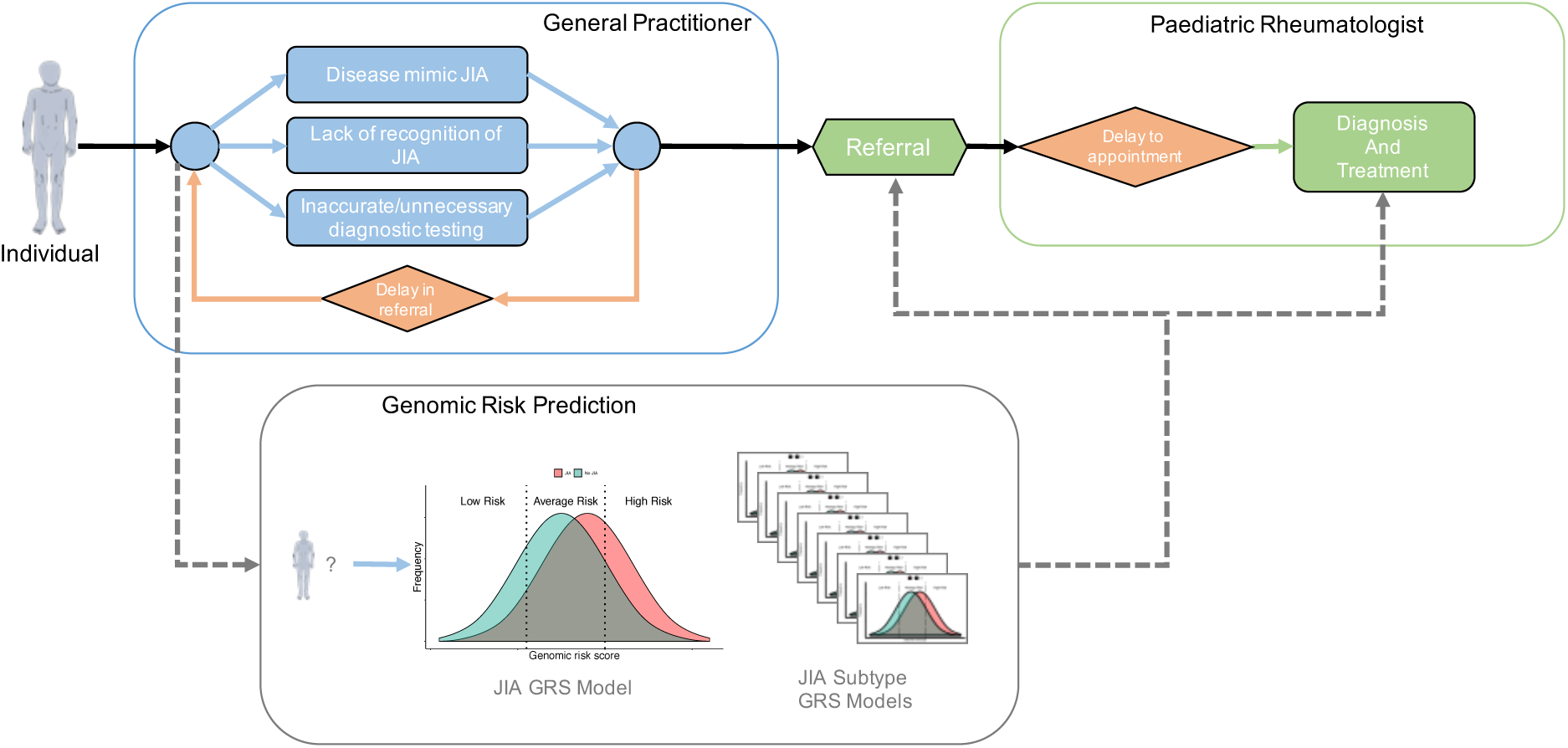
Schematic of typical clinical path from first symptoms to JIA diagnosis and treatment. Potential informative points are included for JIA genomic risk scores to prioritising higher-risk individuals for referral, follow-up, and treatment.

JIA is a complex disease^15^ where susceptibility is due to a complex interaction between genetic and environmental factors. It has been shown that JIA is heritable and that it possesses a genetic architecture similar to other autoimmune diseases, including shared susceptibility genes and strong HLA associations^16–18^. The sibling recurrence risk has been estimated to be 11.6-fold, and for first cousins 5.8-fold^19^. Moreover, a 2015 study estimated SNP-based heritability (SNP-h^2^) of JIA to be ∼75%^18^. Taken together, it is apparent that genetics has an important aetiological role in JIA and may have utility in risk prediction, potentially via stratification of JIA cases from non-cases to aid clinical diagnosis.

Genetics is increasingly used to aid risk prediction, diagnosis and earlier treatment, with HLA testing for various immune disorders being an exemplar. More recently, the clinical utility of genetic and polygenic risk scores for diverse aetiologies, from coeliac disease to cardiovascular diseases, is under intense investigation^20–24^. In coeliac disease, research has shown that a genomic risk score (GRS) based on genome-wide genetic variation (SNPs) can accurately predict coeliac disease cases with high specificity and sensitivity as compared to population-based controls^23^. By using a regularised linear model fitted to individual-level genotype data, it was found that a GRS of ∼200 SNPs provided superior risk prediction over other approaches, including HLA typing. Furthermore, array technologies are relatively affordable with genotyping only needing to be performed once at any point in the lifetime of an individual. GRSs themselves are quantitative measurements of the likelihood that an individual of unknown phenotype has or will have a particular disease, providing potential advantages in terms of flexibility for clinical translation as compared to other tests which are temporally variable or offer only a binary ‘susceptible/not susceptible’ output.

This study aims to create a GRS which in-principle could be used to support the current clinical JIA diagnosis practice. We utilised three large-scale independent cohorts of European ancestry to develop and test a GRS for JIA. Furthermore, we extended the GRS approach to design JIA subtype-specific genetic models, which we externally tested to quantify their potential relative clinical value in supporting each JIA subtype’s time to diagnosis. We make these JIA GRSs publicly available to the community via the Polygenic Score Catalog (www.pgscatalog.org).

## Results

Our analyses utilised three JIA case-control collections from the UK (n=7,505), the Children’s Hospital of Philadelphia (CHOP; n=3,513) and Australian ChiLdhood Arthritis Risk factor Identification sTudY (CLARITY; n=940) which have undergone genome-wide genotyping, quality control, and imputation using standard protocols (for more details see **Methods** and **Table 1**). The UK dataset was used to train the GRS model since it was the dataset with the greatest number of cases and had the least population stratification. For each outcome (JIA or JIA subtype status versus control), GRSs were generated using lasso-penalised regression as implemented in the SparSNP software^25^ and run on the intersecting set of genotypes across the UK, CHOP, and CLARITY datasets (5,545,761 SNPs with MAF>1% across all datasets). After GRS training in the UK dataset, each GRS was externally validated in CHOP and CLARITY (**Figure S1** shows the research workflow).

**Table 1:** Dataset characteristics after imputation and quality control.

|  | Total<br>individuals | SNPs | Number<br>of cases | Number<br>of controls | Number<br>of males | Number<br>of females |
| --- | --- | --- | --- | --- | --- | --- |
| UK | 7,505 | 6,029,891 | 2,324 | 5,181 | 3,433 | 4,072 |
| CHOP | 3,513 | 6,338,131 | 559 | 2,954 | 1,671 | 1,842 |
| CLARITY | 940 | 5,743,016 | 362 | 578 | 460 | 480 |

### Training and external validation of the JIA genomic risk score

We used 10×10-fold cross-validation to tune the penalised model and estimate the area under the receiver operating characteristic curve (AUC) in the UK dataset, achieving a maximum AUC=0.671 (95% CI 0.668−0.674) and selecting 26 SNPs in the model (**Figure S2**). The selected SNPs and their weights in the GWAS clearly showed the strong HLA dependency of JIA (**Figure S3)**. The final model (SNP weights) was held fixed for external validation GRS (http://www.pgscatalog.org/pgs/PGS000114/).

We applied the GRS to the CLARITY and CHOP datasets and evaluated the prediction power of our model in terms of AUC and odds ratios (**Table *2***). Overall, the GRS showed highly consistent performance across both validation datasets, with AUCs of 0.657 and 0.671 for CHOP and CLARITY, respectively. The corresponding odds ratios per s.d. of GRS were 1.831 (95% CI 1.685-1.991) and 2.008 (95% CI 1.731-2.345) in CHOP and CLARITY, respectively. The associations of the GRS with JIA status were unattenuated when adjusting for the top 10 genetic principal components (PCs) and sex. Furthermore, we compared the GRS model with a model generated using known SNPs highly association with JIA (**Table 1** from Hinks et al.^16^). Using this model, we achieved AUC of 0.614, 0.642 and 0.648 for UK, CHOP, and CLARITY respectively, showing that the GRS model was a stronger risk predictor.

**Table 2:** The predictive power of the GRS in the validation sets. Based on logistic regression on the test sets, optionally adjusted for sex and 10 genetic principal components. Effect sizes are per standard deviation of the GRS.

|  | AUC (95% CI) | OR (95% CI) |
| --- | --- | --- |
| **CHOP** |  |  |
| Sex+PCs | 0.677 (0.654-0.701) |  |
| GRS | 0.657 (0.631-0.683) | 1.831 (1.685-1.991) |
| GRS+Sex+PCs | 0.735 (0.712-0.758) | 1.838 (1.686-2.007) |
| **CLARITY** |  |  |
| Sex+PCs | 0.671 (0.636-0.706) |  |
| GRS | 0.671 (0.635-0.706) | 2.008 (1.731-2.345) |
| GRS+Sex+PCs | 0.738 (0.705-0.770) | 2.085 (1.773-2.471) |

Recent work has also shown that a metaGRS approach can substantially improve genomic prediction of common diseases^21^, thus we compared the metaGRS method to the lasso-penalised regression above (**Supplementary Material**). We did not observe that a JIA metaGRS, based on a set of related autoimmune disease GWAS summary statistics, significantly improved upon the JIA GRS above, thus subsequent analyses utilised lasso-regression.

### Subtype analysis

We next extended our analysis to consider subtypes of JIA and construct subtype-specific GRSs thereof. The International League of Associations of Rheumatology (ILAR) classification system recognises seven subtypes of JIA, systemic arthritis, oligoarthritis, rheumatoid-factor-positive polyarthritis (RF-Positive), rheumatoid-factor-negative polyarthritis (RF-Negative), enthesitis-related arthritis (ERA), psoriatic arthritis, and undifferentiated arthritis^5^. Subtypes vary substantially in average age at onset, sex distribution, number of joint affected, and clinical features (see **Figure *2***)^26^. Of particular interest clinically is the time between onset of symptoms to visiting a paediatric rheumatologist which has been estimated to vary from 11 months (range: 8 to 70 weeks) in the case of enthesitis-related JIA to 1 month (range: 2 to 36 weeks) for systemic JIA^14,27^. The heterogeneity of JIA was reflected in our case data; all seven subtypes were present, albeit at frequencies ranging from common (∼41% of UK cases were oligoarthritis JIA) to relatively rare (∼4% of UK cases were undifferentiated JIA) (**Table *3***).

**Table 3:** Characteristics of JIA subtypes across cohorts, including rate (%) of each subtype amongst cases of each cohort. CLARITY excludes 29 cases with no subtype classification.

|  | UK (2,324 cases) |  |  | CHOP (559 cases) |  |  | CLARITY (333 cases) |  |  |
| --- | --- | --- | --- | --- | --- | --- | --- | --- | --- |
|  | Rate (%) | Males | Females | Rate (%) | Males | Females | Rate (%) | Males | Females |
| **Enthesitis-related** | 7.5 | 119 | 56 | 11.8 | 40 | 26 | 4.4 | 13 | 3 |
| **Oligoarthritis** | 41.4 | 301 | 660 | 36.3 | 39 | 164 | 43.9 | 42 | 117 |
| **RF-Negative** | 22 | 144 | 367 | 24.2 | 34 | 101 | 20.7 | 23 | 52 |
| **RF-Positive** | 8.1 | 34 | 155 | 5.2 | 1 | 28 | 3 | 1 | 10 |
| **Psoriatic** | 5.9 | 50 | 87 | 7.2 | 11 | 29 | 5 | 12 | 6 |
| **Undifferentiated** | 3.5 | 41 | 41 | 4.7 | 8 | 18 | 7.5 | 13 | 14 |
| **Systemic** | 11.6 | 125 | 144 | 10.6 | 24 | 36 | 7.5 | 12 | 15 |

For each of the seven subtypes, we utilised the UK dataset to train subtype specific GRSs, employing a similar lasso-regression in cross-validation approach as for the JIA GRS above (see **Methods**). Each subtype GRS was trained on that subtype’s cases and all non-JIA controls, with all other JIA subtypes were excluded from training, and then externally validated in CHOP and CLARITY. We found that there was a high degree of variability in discrimination between subtype GRSs, with some subtypes displaying cross-validated AUCs greater than the JIA GRS and others not being significantly different than random chance (AUC=0.5; **Figure 2**).

**Figure 2:**
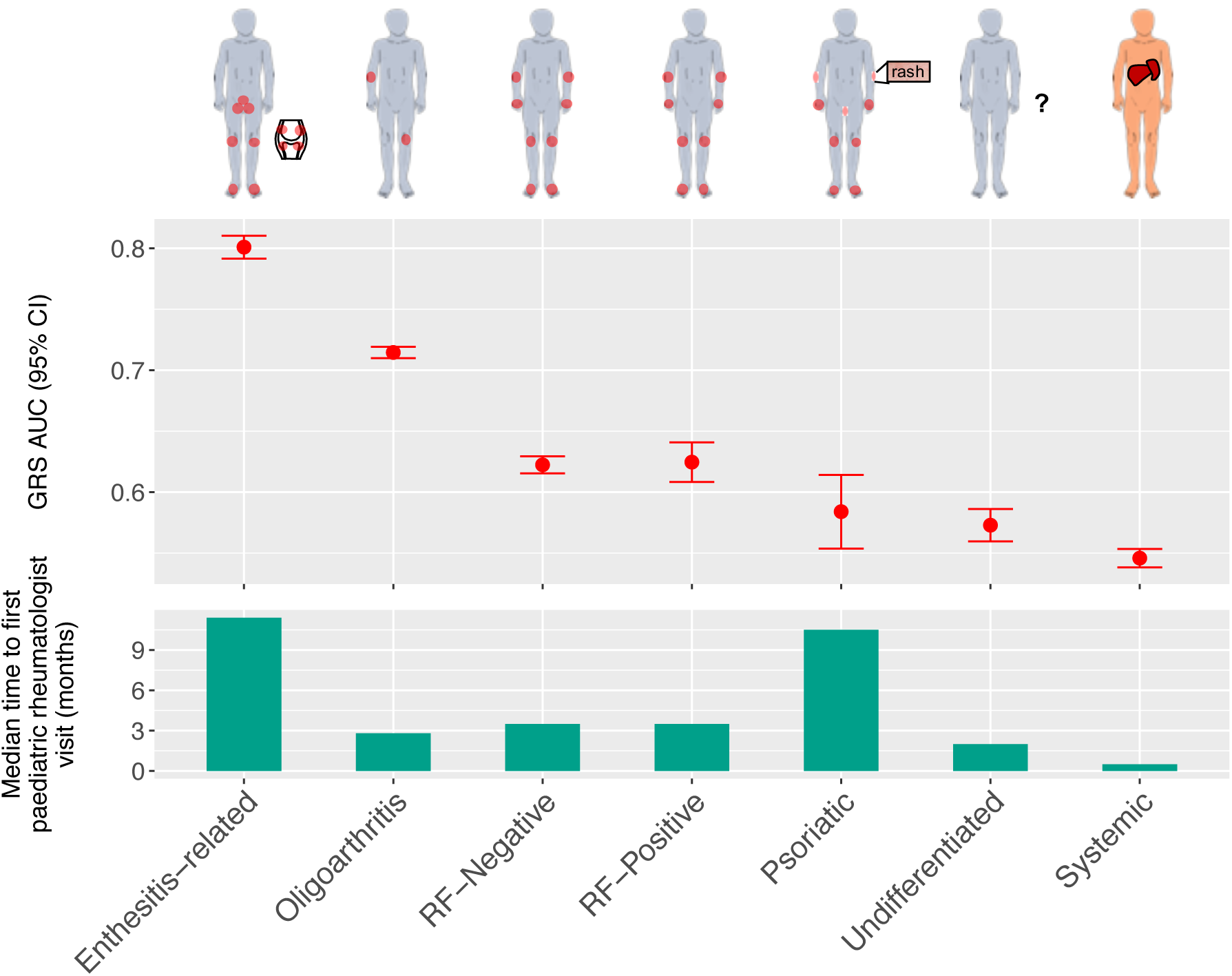
10×10 cross-validated AUC (LOESS-smoothed) AUC achieved by training the seven JIA subtype specific models (top) and median time taken by an individual with JIA to be referred for first time to a paediatric rheumatologist visit (in months; bottom)^27^.

When compared to a previous estimate of the median time for a child with JIA to be referred to a paediatric rheumatologist visit^27^, we found that the JIA subtype (enthesitis-related) with the longest time-to-referral (∼11 months) also had the strongest subtype GRS (AUC=0.80 with 149 SNPs). This result may be partially due to circulating HLA-B27 antigen being a component of the enthesitis-related subtype classification criteria^5,28^. To test this, we utilised previously reported *HLA-B27* tag SNPs^29,30^ (**Supplementary Material**), and found that our enthesitis-related JIA GRS still outperformed the *HLA-B27* tag SNPs model in the UK dataset (AUC=0.75). The next strongest subtype GRS (AUC=0.71 with 25 SNPs) was for oligoarthritis JIA, which has a median time-to-referral of ∼3 months. While JIA subtype rates are not well characterised, oligoarthritis JIA is the most common clinical subtype reported^31^.

The weakest subtype GRSs were for the undifferentiated (AUC=0.573 with 119 SNPs) and systemic (AUC=0.546 with 3,430 SNPs) subtypes. This was not unexpected as these subtypes are somewhat different to the other five subtypes. Systemic JIA is considered an autoinflammatory disease and, while there is little genetic overlap with the other JIA subtypes^32^, it has strong genetic signals from the HLA class II molecule encoded by *HLA-DRB1*11*, confirming the role of the adaptive immune system^33^. Children are diagnosed with the undifferentiated subtype when their symptoms do not fit within other subtypes, or meet the criteria for multiple subtypes.

External validation of the subtype GRSs in CLARITY showed highly consistent AUC estimates with cross-validation performance in the UK (**Table *4***). The CHOP dataset showed somewhat less consistent external validation than CLARITY for subtype GRSs, but in both CLARITY and CHOP the top performing subtype GRSs were both the enthesitis-related JIA GRS (AUCs of 0.83 and 0.69, respectively) and oligoarthritis JIA GRS (0.75 and 0.77, respectively). In both external validations, the systemic JIA subtype GRS had poor predictive capacity with AUCs of 0.47 and 0.55 in CLARITY and CHOP, respectively.

**Table 4:**
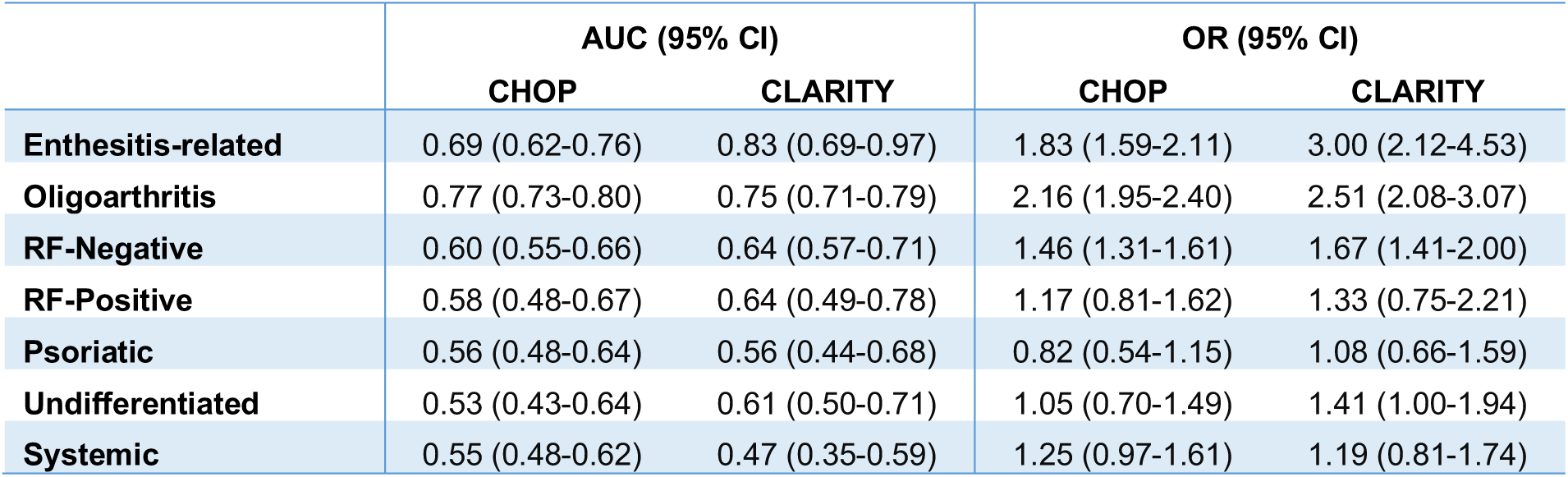
External validation of the subtype-specific GRSs in CLARITY and CHOP. Based on logistic regression on the test sets. Effect sizes are per standard deviation of the GRS.

|  | AUC (95% CI) |  | OR (95% CI) |  |
| --- | --- | --- | --- | --- |
|  | CHOP | CLARITY | CHOP | CLARITY |
| **Enthesitis-related** | 0.69 (0.62-0.76) | 0.83 (0.69-0.97) | 1.83 (1.59-2.11) | 3.00 (2.12-4.53) |
| **Oligoarthritis** | 0.77 (0.73-0.80) | 0.75 (0.71-0.79) | 2.16 (1.95-2.40) | 2.51 (2.08-3.07) |
| **RF-Negative** | 0.60 (0.55-0.66) | 0.64 (0.57-0.71) | 1.46 (1.31-1.61) | 1.67 (1.41-2.00) |
| **RF-Positive** | 0.58 (0.48-0.67) | 0.64 (0.49-0.78) | 1.17 (0.81-1.62) | 1.33 (0.75-2.21) |
| **Psoriatic** | 0.56 (0.48-0.64) | 0.56 (0.44-0.68) | 0.82 (0.54-1.15) | 1.08 (0.66-1.59) |
| **Undifferentiated** | 0.53 (0.43-0.64) | 0.61 (0.50-0.71) | 1.05 (0.70-1.49) | 1.41 (1.00-1.94) |
| **Systemic** | 0.55 (0.48-0.62) | 0.47 (0.35-0.59) | 1.25 (0.97-1.61) | 1.19 (0.81-1.74) |

## Discussion

The diagnosis of juvenile idiopathic arthritis has a pressing, yet unmet, clinical need. In this study, we aimed to address the paucity of molecular tools to aid the entirely clinical diagnosis of JIA by leveraging the wealth of human genomic data developed over the last decade to develop a series of genomic risk scores for JIA. We showed that genomic machine learning can yield highly predictive GRSs for JIA as a composite diagnosis as well as subtype-specific GRSs which can classify for example the common oligoarthritis JIA subtype as well as the enthesitis-related JIA subtype, which can present with non-specific pains initially and be difficult to diagnose clinically. Given the cost effectiveness of a genotyping array and the time invariant properties of assaying germline DNA, these JIA GRSs hold promise for rapid clinical translation with at-risk children being non-invasively stratified as high risk much earlier in the diagnostic pathway. It also allows for children with low risk non-inflammatory disease to be appropriately triaged and managed earlier. To facilitate translation and clinical uptake, we have made the genetic variants and weights of the JIA GRSs developed here freely available (http://www.pgscatalog.org/pgs/PGS000114/).

A strength of this study was that the JIA GRS was developed on a UK dataset and externally validated in two independent studies in Australia and the US, indicating the robustness of the score. A limitation of the current study is that the vast majority of participants in our cohorts (and the original JIA GWASs) were of European descent and we were unable to assess the performance of a JIA GRS in individuals of non-European ancestry^34^.

Many clinicians at the primary health level, from general practitioners through to tertiary hospital settings, often have difficulty recognising and diagnosing JIA in children and adolescents. There are many non-inflammatory conditions that are common in children and other conditions presenting with musculoskeletal pain that mimic JIA. In primary care, accessing paediatric rheumatology specialist services is difficult as wait lists are usually lengthy and access to care is problematic due to workforce shortages worldwide^35^.

Therefore, in the diagnostic algorithm a JIA-GRS would be particularly useful when assessing children with musculoskeletal symptoms who may be JIA cases, for example enabling the general practitioner to facilitate appropriate triage to a paediatric rheumatologist, and thus reduce pain, complications of the disease and poor long-term health outcomes.

## Methods

### Ethics Statement

All participants gave informed consent and the study protocols were approved by the relevant institutional or national ethics committees. The Australian CLARITY cohort collection was approved by the Royal Children’s Hospital Human Research Ethics Committee; UK ethical approval was obtained from the North West Multi-centre for Research Ethics Committee (MREC:02/8/104 and MREC:99/8/84), West Midlands Multi-centre Research Ethics Committee (MREC:02/7/106), North West Research Ethics Committee (REC:09/H1008/137), and the NHS Research Ethics Committee (REC:05/Q0508/95); and the US CHOP cohort collection was approved by the institutional review boards of the Texas Scottish Rite Hospital for Children, the Children’s Mercy Hospitals and Clinics, and the Children’s Hospital of Philadelphia.

### Phenotype and clinical data

The International League of Associations for Rheumatology (ILAR) classification system^5^ provides generally accepted guidelines for researchers and clinicians to delineate the seven mutually-exclusive categories of JIA based on the predominate clinical and laboratory features. For example, to diagnose the enthesitis-related JIA subtype, an individual has have at least two of the following conditions: presence of HLA-B27 antigen; onset of arthritis in a male over age six; acute anterior uveitis; presence of or history of sacroiliac joint tenderness and/or inflammatory lumbosacral pain; or have a first-degree relative with history of ankylosing spondylitis, enthesitis-related arthritis, Reiter’s syndrome, or acute anterior uveitis.

In this work, the JIA diagnosis for the three cohorts was made according to the ILAR revised criteria and all JIA cases were of age of onset <16 years old. In the UK cohort^16^, the JIA cases were obtained from five sources: The British Society for Paediatric and Adolescent Rheumatology National Repository of JIA; a group of UK cases with long-standing JIA described previously^36^; a cohort collected as part of the Childhood Arthritis Prospective Study^10^; a cohort of children recruited for the SPARKS-CHARM (Childhood Arthritis Response to Medication)^37^; and an ongoing collection of UK cases from the UK JIA Genetics Consortium. The UK controls were from the shared UK 1958 Birth cohort and UK Blood Services Common Controls, established as part of the WTCCC^38^.

For the CHOP cohort^18,39^, the clinical data of JIA cases were collected from the electronic health records (EHR) completed by the pediatric rheumatology specialist within the Division of Rheumatology at CHOP and abstracted into a JIA Registry maintained within the Center for Applied Genomics (CAG) at CHOP. Control subjects were unrelated and disease-free children recruited by the CAG recruitment team within the CHOP Healthcare Network. In addition, control subjects had no history of JIA or other chronic illnesses and were screened as negative for a diagnosis of autoimmune diseases, based on data from CHOP’s electronic health record and by intake questionnaires obtained by the recruiting staff from the CHOP Center for Applied Genomics.

Finally, in the CLARITY cohorts^13,40^, all the cases were aged 0–18 years at interview, with diagnosis of JIA by a paediatric rheumatologist. Incident cases were defined as children recruited within six months of diagnosis and prevalent cases were defined as those children diagnosed more than 6 months before recruitment and since 1997. Controls were recruited through the Royal Children’s Hospital Day Surgery Unit. Exclusion criteria for cases and controls were the presence of major congenital abnormalities or illness that would forgo school attendance in the one year prior to recruitment.

### Genotype data and quality control

All collected cases are pediatric rheumatologist-diagnosed and controls have no history of any autoimmune disease. Additionally, all participants are white European descent with the exception of ∼20% of the participants in CLARITY, who are of non-European ancestries. Finally, all genotyped included in each dataset were aligned to the GRCh37/hg19 assembly build and passed stringent quality control measures (detailed below).

The initial UK dataset consists of 2,758 cases and 5,187 controls. Controls were obtained from the Wellcome Trust Case Control Consortium (WTCCC) which have been demonstrated to be well-matched to UK JIA cases^16,41^. UK cases were collected through five cohort studies recruiting from multiple tertiary centers around the UK, where 1,670 were genotyped on the Illumina HumanOmniExpress array and 1,088 on the Illumina HumanCoreExome array. The CHOP (US) dataset consisted of 1,229 cases from US and Norway, and 5,512 pediatric controls, all recruited from within the CHOP Healthcare Network. All CHOP controls and 1,008 JIA cases were genotyped using Illumina HumanHap550 or Human610-Quad arrays^40^ and the remaining 250 cases were genotyped using the Illumina HumanCore array. Lastly, the Australian CLARITY (ChiLdhood Arthritis Risk factor Identification sTudY)^13^ consisted of 558 cases and 704 controls collected from the Royal Children’s Hospital and Monash Medical Centre in Melbourne. All controls and 406 cases were genotyped using the Illumina HumanCore array, with the remaining 152 cases genotyped on the Illumina HumanHap550^17,18^.

Before genotype imputation, we applied stringent quality control (QC) procedures standardised across all datasets. The initial QC performed, using plink1.9^42,43^, consisted of removing non-autosomal SNPs, SNPs with MAF<1%, SNPs and individuals with missingness>10%, and SNPs with deviations from Hardy-Weinberg Equilibrium in controls (P<10^−3^). Additionally, using KING v2.1.5^44^, we identified and removed one of two individuals with a second or higher degree relatedness within the datasets. The resulting datasets consisted of 7,505 individuals (2,324 cases and 5,181 controls) with 144,964 SNPs for UK, and 6,741 individuals (1,229 cases and 5,512 controls) with 489,672 SNPs for CHOP. The CLARITY dataset consisted of three batches; after quality control, these consisted of 152 individuals (all cases) with 490,629 SNPs, 838 individuals (247 cases and 591 controls) with 246,814 SNPs, and 118 individuals (5 cases and 113 controls) with 243,662 SNPs, respectively.

For genotype imputation, we used the Michigan Server to impute the datasets using Minimac3 and the 2016 Haplotype Reference Consortium (HRC) as the reference panel^45^. We merged all the subsets of CLARITY into a single dataset. Then, for each dataset (UK, CHOP and CLARITY), we removed multi-allelic and duplicated SNPs, SNPs with imputation *r*^2^<0.5 and MAF<0.01, SNPs deviating from the Hardy-Weinberg Equilibrium in controls (P<10^−3^), and those with ambiguous strand (A/T or C/G alleles).

Next, we performed principal component analysis (PCA) using FlashPCA2^46,47^ over the filtered data (**Figures S8**–**S11)**. For each dataset we selected the largest homogeneous subset of individuals based on the first five principal components within each set, where we defined as outliers the set of cases (controls) individuals that clearly differed in the PCA plots from the controls (cases). The UK dataset was kept in its entirety; for CLARITY n=168 individuals were removed and for CHOP n=3,228 individuals were removed (including non-US individuals). **Figures S12** and **S13** show the principal component analysis for the resulting subsets. **Table 1** shows the final datasets; for the analysis we used the n=5,545,761 genotyped and imputed SNPs which were available post-QC on all three datasets.

Before training our model we also estimated the heritability and SNP-heritability of JIA in our datasets. The sibling recurrence risk of JIA has previously been estimated to be 11.6-fold^19^, and for first cousins 5.8-fold; assuming the JIA prevalence to be 1 in 1000, we estimated JIA heritability to be approximately 54%. For SNP-heritability, we used GCTA version 1.91.7^27,48^ and adjusted for the first ten principal components together with an assumed JIA population prevalence of 1/1000. The estimated SNP-heritability was 0.25 (std err 0.02) for UK, 0.37 (std err 0.13) for CLARITY, and 0.51 (std err 0.07) for CHOP.

### Development and validation of the genomic risk score

Because it was the most homogenous dataset with the largest case sample size (2,324 cases, 5,181 controls), we utilised the UK dataset to train our models. To account for potential confounding by the case/control genotyping batch in the UK dataset, we used logistic regression of case/control status on sex and the first 10 genetic PCs. The PCs were computed over a subset of the SNPs of UK, excluding the HLA region as well as known or putative JIA risk loci^16^ (defined here as SNPs with P<10^−5^ and all SNPs within 1 Mb of the former). The residuals from the regression were then used as the phenotype for constructing the GRS.

To create the GRS we used SparSNP^25^, which is an efficient implementation of a lasso-penalised linear model which has been previously shown to outperform other methods when there are known to be strong effects within regions of complex LD, such as the MHC^49^. This model is induced by minimising the L1-penalised squared loss function (standard lasso) over *N* individuals and *m* SNPs,

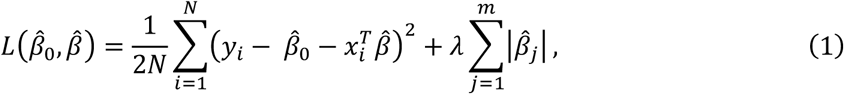

where *x*_*i*_ is the *m*-vector of genotypes for the *i*th sample in allele-dosage coding {0,1,2}, *y*_*i*_ is the phenotype, 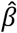 is the *m*-vector of weights, 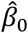 is the intercept, and *λ* is the L1 penalty. All SNPs in the selected training dataset are considered in the model but typically only a small number of SNPs receive a non-zero weight. The number of SNPs with non-zero weight varies depending on the value of the L1 penalty (higher penalties lead to fewer SNPs) which was tuned via 10-fold cross-validation. The optimal number of SNPs selected (non-zero weighted) in the chosen model is decided based on the model with the highest average AUC (or *R*^2^ if the phenotype is continuous) across the replications. We also explored adding an L2 penalty to the model (elastic-net); however, no significant improvement was found.

Once a model was chosen, we computed genomic risk scores (GRS) for each of the test datasets (CHOP and CLARITY). Assuming that the number of SNPs is *m*, then the GRS *g*_*i*_ for a new individual with genotypes *x*_*i*_ is

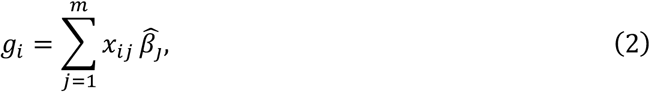

where and 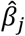 are the SNP weights obtained from the model. Subsequently, in order to validate the GRS, we evaluated it using logistic regression in the CHOP and CLARITY datasets, adjusting for sex and ten first genetic principal components of each test set.

## Data Availability

For the individual-level data, note that CHOP is available through the eMERGE Network dbGaP and the WTCCC controls are available through the Wellcome Trust Case Control Consortium webpage https://www.wtccc.org.uk/). For the individual-level genotype data for the UK JIA cases and CLARITY, researchers should contact the cohort PIs (Prof Wendy Thomson and Dr Jane Munro, respectively). Finally, the JIA GRSs presented will be publicly available to the community via the Polygenic Score Catalog (www.pgscatalog.org)

https://www.wtccc.org.uk/

## Acknowledgments

This study was supported in part by the Victorian Government’s OIS Program, the Australian National Health and Medical Research Council (NHMRC Project no. 1122744), the Murdoch Children’s Research Institute, and the Royal Children’s Hospital Foundation (grant no. 2017-896). Also, this work was supported by core funding from the UK Medical Research Council (MR/L003120/1), the British Heart Foundation (RG/13/13/30194; RG/18/13/33946) and the National Institute for Health Research (Cambridge Biomedical Research Centre at the Cambridge University Hospitals NHS Foundation Trust)*. It was also supported by Health Data Research UK, which is funded by the UK Medical Research Council, Engineering and Physical Sciences Research Council, Economic and Social Research Council, Department of Health and Social Care (England), Chief Scientist Office of the Scottish Government Health and Social Care Directorates, Health and Social Care Research and Development Division (Welsh Government), Public Health Agency (Northern Ireland), British Heart Foundation and Wellcome. GA was supported by an NHMRC Early Career Fellowship (no. 1090462). MI was supported by the Munz Chair of Cardiovascular Prediction and Prevention. This study acknowledges the use of the following UK JIA cohort collections: The Biologics for Children with Rheumatic Diseases (BCRD) study (funded by Arthritis Research UK Grant 20747). The British Society for Paediatric and Adolescent Rheumatology Etanercept Cohort Study (BSPAR-ETN) (funded by a research grant from the British Society for Rheumatology (BSR). BSR has previously also received restricted income from Pfizer to fund this project). Childhood Arthritis Prospective Study (CAPS) (funded by Versus Arthritis, grant reference number 20542), Childhood Arthritis Response to Medication Study (CHARMS) (funded by Sparks UK, reference 08ICH09, and the Medical Research Council, reference MR/M004600/1), United Kingdom Juvenile Idiopathic Arthritis Genetics Consortium (UKJIAGC). Genotyping of the UK JIA case samples were supported by the Versus Arthritis grants reference numbers 20385 and 21754. This research was funded by the NIHR Manchester Biomedical Research Centre and supported by the Manchester Academic Health Sciences Centre (MAHSC). The views expressed are those of the author(s) and not necessarily those of the NHS, the NIHR or the Department of Health. We would like to acknowledge the assistance given by IT Services and the use of the Computational Shared Facility at The University of Manchester. Finally, the CHOP data used were funded by an Institute Development Fund to the CAG center from The Children’s Hospital of Philadelphia and by NIH grant, U01-HG006830, from the NHGRI-sponsored eMERGE Network.

## Footnotes

* The views expressed are those of the authors and not necessarily those of the NHS, the NIHR or the Department of Health and Social Care.

## References

1. Ravelli, A. & Martini, A. Juvenile idiopathic arthritis. Lancet vol. 369 767–778 (2007).

2. Manners, P. J. & Diepeveen, D. A. Prevalence of juvenile chronic arthritis in a population of 12-year-old children in urban Australia. Pediatrics 98, 84–90 (1996).

3. Szer, I. S., Kimura, Y., Malleson, P. & Southwood, T. Arthritis in children and adolescents : juvenile idiopathic arthritis. (Oxford University Press, 2006).

4. Symmons, D. P. et al. Pediatric rheumatology in the United Kingdom: data from the British Pediatric Rheumatology Group National Diagnostic Register. J. Rheumatol. 23, 1975–80 (1996).

5. Petty, R. E. et al. International League of Associations for Rheumatology Classification of Juvenile Idiopathic Arthritis: Second Revision, Edmonton, 2001. Journal of Rheumatology vol. 31 390–392 (2004).

6. Albers, H. M. et al. Time to treatment as an important factor for the response to methotrexate in juvenile idiopathic arthritis. Arthritis Care Res. 61, 46–51 (2009).

7. Hinze, C., Gohar, F. & Foell, D. Management of juvenile idiopathic arthritis: Hitting the target. Nature Reviews Rheumatology vol. 11 290–300 (2015).

8. Van Dijkhuizen, E. H. P. & Wulffraat, N. M. Early predictors of prognosis in juvenile idiopathic arthritis: A systematic literature review. Annals of the Rheumatic Diseases vol. 74 1996–2005 (2015).

9. Wallace, C. A. et al. Trial of early aggressive therapy in polyarticular juvenile idiopathic arthritis. Arthritis Rheum. 64, 2012–2021 (2012).

10. Adib, N. et al. Association between duration of symptoms and severity of disease at first presentation to paediatric rheumatology: Results from the Childhood Arthritis Prospective Study. Rheumatology 47, 991–995 (2008).

11. Barber, C. E. H. et al. Testing population-based performance measures identifies gaps in juvenile idiopathic arthritis (JIA) care. BMC Health Serv. Res. 19, 572 (2019).

12. McErlane, F. et al. Trends in paediatric rheumatology referral times and disease activity indices over a ten-year period among children and young people with Juvenile Idiopathic Arthritis: Results from the childhood arthritis prospective Study. Rheumatol. (United Kingdom) 55, 1225–1234 (2016).

13. Ellis, J. A. et al. CLARITY - ChiLdhood Arthritis Risk factor Identification sTudY. Pediatr. Rheumatol. 10, (2012).

14. Foster, H. E. et al. Delay in access to appropriate care for children presenting with musculoskeletal symptoms and ultimately diagnosed with juvenile idiopathic arthritis. Arthritis Care Res. 57, 921–927 (2007).

15. Glass, D. N. & Giannini, E. H. Juvenile rheumatoid arthritis as a complex genetic trait. Arthritis and Rheumatism vol. 42 2261–2268 (1999).

16. Hinks, A. et al. Dense genotyping of immune-related disease regions identifies 14 new susceptibility loci for juvenile idiopathic arthritis. Nat. Genet. 45, 664–669 (2013).

17. Li, Y. R. et al. Meta-analysis of shared genetic architecture across ten pediatric autoimmune diseases. Nat. Med. 21, 1018–1027 (2015).

18. Li, Y. R. et al. Genetic sharing and heritability of paediatric age of onset autoimmune diseases. Nat. Commun. 6, (2015).

19. Prahalad, S. et al. Quantification of the familial contribution to juvenile idiopathic arthritis. Arthritis Rheum. 62, 2525–2529 (2010).

20. Abraham, G. & Inouye, M. Genomic risk prediction of complex human disease and its clinical application. Current Opinion in Genetics and Development vol. 33 10–16 (2015).

21. Inouye, M. et al. Genomic Risk Prediction of Coronary Artery Disease in 480,000 Adults: Implications for Primary Prevention. J. Am. Coll. Cardiol. 72, 1883–1893 (2018).

22. Lambert, S. A., Abraham, G. & Inouye, M. Towards clinical utility of polygenic risk scores. Hum. Mol. Genet. 28, R133–R142 (2019).

23. Abraham, G. et al. Accurate and Robust Genomic Prediction of Celiac Disease Using Statistical Learning. PLoS Genet. 10, e1004137 (2014).

24. Sharp, S. A. et al. Development and standardization of an improved type 1 diabetes genetic risk score for use in newborn screening and incident diagnosis. in Diabetes Care vol. 42 200–207 (American Diabetes Association Inc., 2019).

25. Abraham, G., Kowalczyk, A., Zobel, J. & Inouye, M. SparSNP: Fast and memory-efficient analysis of all SNPs for phenotype prediction. BMC Bioinformatics 13, 88 (2012).

26. Hinks, A. et al. Fine-mapping the MHC locus in juvenile idiopathic arthritis (JIA) reveals genetic heterogeneity corresponding to distinct adult inflammatory arthritic diseases. Ann. Rheum. Dis. 76, 765–772 (2017).

27. Freychet, C. et al. Medical pathways of children with juvenile idiopathic arthritis before referral to pediatric rheumatology centers. Jt. Bone Spine 86, 739–745 (2019).

28. Berntson, L. et al. HLA-B27 predicts a more extended disease with increasing age at onset in boys with Juvenile idiopathic arthritis. J. Rheumatol. 35, 2055–2061 (2008).

29. Evans, D. M. et al. Interaction between ERAP1 and HLA-B27 in ankylosing spondylitis implicates peptide handling in the mechanism for HLA-B27 in disease susceptibility. Nat. Genet. 43, 761–767 (2011).

30. Cortes, A. et al. Identification of multiple risk variants for ankylosing spondylitis through high-density genotyping of immune-related loci. Nat. Genet. 45, 730–738 (2013).

31. Guzman, J. et al. The outcomes of juvenile idiopathic arthritis in children managed with contemporary treatments: Results from the reacch-out cohort. Ann. Rheum. Dis. 74, 1854–1860 (2015).

32. Ombrello, M. J. et al. Genetic architecture distinguishes systemic juvenile idiopathic arthritis from other forms of juvenile idiopathic arthritis: Clinical and therapeutic implications. Ann. Rheum. Dis. 76, (2017).

33. Ombrello, M. J. et al. HLA-DRB1*11 and variants of the MHC class II locus are strong risk factors for systemic juvenile idiopathic arthritis. Proc. Natl. Acad. Sci. U. S. A. 112, 15970–15975 (2015).

34. Martin, A. R. et al. Clinical use of current polygenic risk scores may exacerbate health disparities. Nat. Genet. 51, 584–591 (2019).

35. Henrickson, M. Policy challenges for the pediatric rheumatology workforce: Part III. the international situation. Pediatr. Rheumatol. 9, 26 (2011).

36. Packham, J. C. Long-term follow-up of 246 adults with juvenile idiopathic arthritis: functional outcome. Rheumatology 41, 1428–1435 (2002).

37. Moncrieffe, H. et al. Generation of novel pharmacogenomic candidates in response to methotrexate in juvenile idiopathic arthritis: Correlation between gene expression and genotype. Pharmacogenet. Genomics 20, 665–676 (2010).

38. Burton, P. R. et al. Genome-wide association study of 14,000 cases of seven common diseases and 3,000 shared controls. Nature 447, 661–678 (2007).

39. Finkel, T. H. et al. Variants in CXCR4 associate with juvenile idiopathic arthritis susceptibility. BMC Med. Genet. 17, 24 (2016).

40. Ellis, J. A. et al. Epistasis amongst PTPN2 and genes of the vitamin D pathway contributes to risk of juvenile idiopathic arthritis. J. Steroid Biochem. Mol. Biol. 145, 113–120 (2015).

41. Cobb, J. et al. Genome-wide data reveal novel genes for methotrexate response in a large cohort of juvenile idiopathic arthritis cases. Pharmacogenomics J. 14, 356–364 (2014).

42. Chang, C. C. et al. Second-generation PLINK: Rising to the challenge of larger and richer datasets. Gigascience 4, (2015).

43. Purcell, S. et al. PLINK: A tool set for whole-genome association and population-based linkage analyses. Am. J. Hum. Genet. 81, 559–575 (2007).

44. Manichaikul, A. et al. Robust relationship inference in genome-wide association studies. Bioinformatics 26, 2867–2873 (2010).

45. Das, S. et al. Next-generation genotype imputation service and methods. Nat. Genet. 48, 1284–1287 (2016).

46. Abraham, G. & Inouye, M. Fast principal component analysis of large-scale genome-wide data. PLoS One 9, (2014).

47. Abraham, G., Qiu, Y. & Inouye, M. FlashPCA2: principal component analysis of Biobank-scale genotype datasets. Bioinformatics 33, 2776–2778 (2017).

48. Yang, J., Lee, S. H., Goddard, M. E. & Visscher, P. M. GCTA: A tool for genome-wide complex trait analysis. Am. J. Hum. Genet. 88, 76–82 (2011).

49. Abraham, G., Kowalczyk, A., Zobel, J. & Inouye, M. Performance and Robustness of Penalized and Unpenalized Methods for Genetic Prediction of Complex Human Disease. Genet. Epidemiol. 37, 184–195 (2013).

